## Supplementary 1 for "An analysis of NHS 111 demand for primary care services"

### Symptom Groups

The study data collection period (January to December, 2021) coincided with the third English lockdown due to COVID-19. While several symptom group weekly frequencies did not change, for example pain on passing urine, others, particularly those which might be exacerbated by the relaxing of COVID-19 restrictions, for example coughs and sore throats, did see an increase.

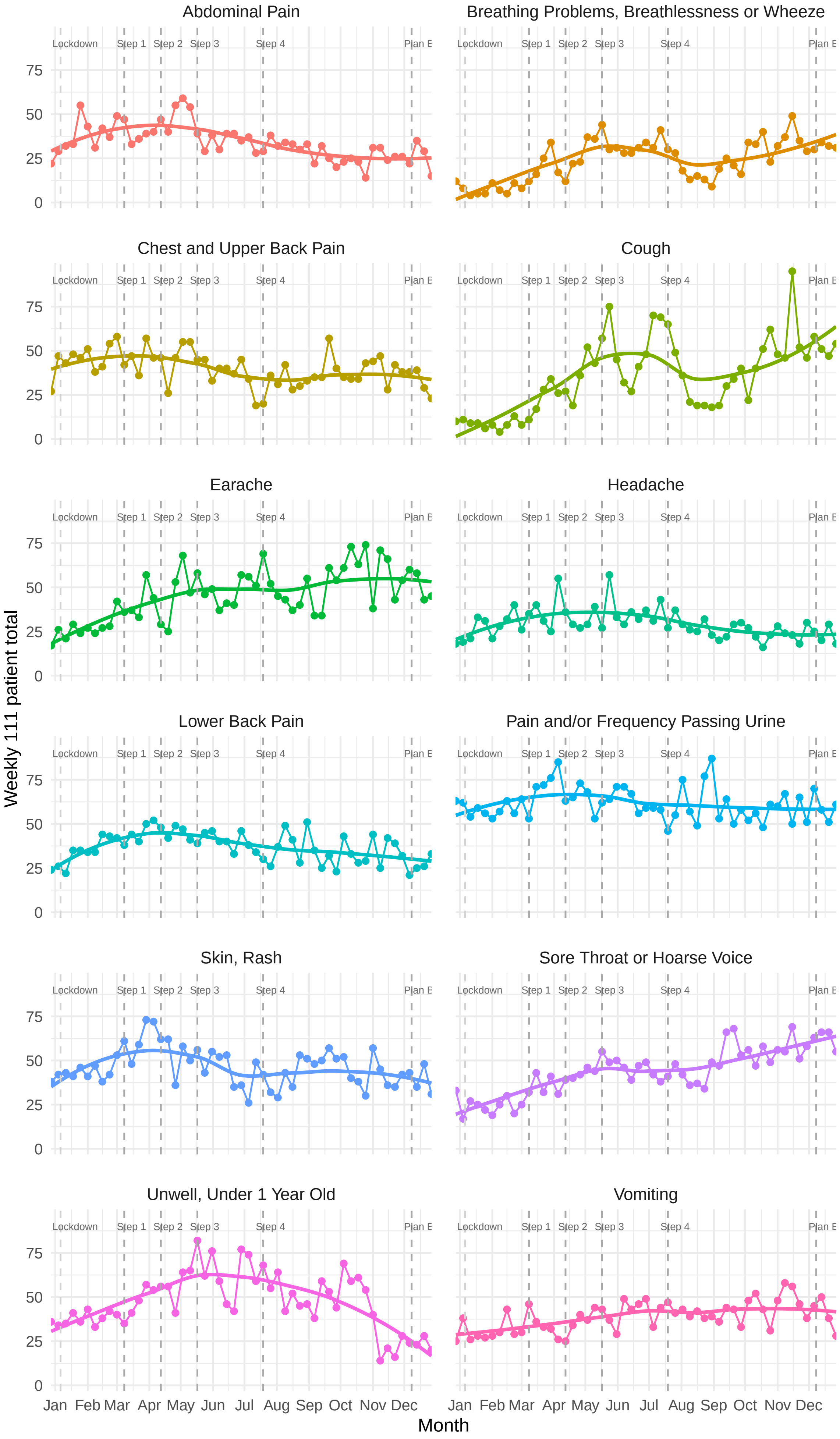


Figure 1: Top 12 weekly 111 symptom groups allocated to callers
