## Supplementary 2 for "An analysis of NHS 111 demand for primary care services"

### Age distribution of callers to NHS 111

The age distribution was bimodal, with peaks seen in patients less than a year old, and in patients aged between 20–30 years. Callers were more commonly female across virtually the entire age range.

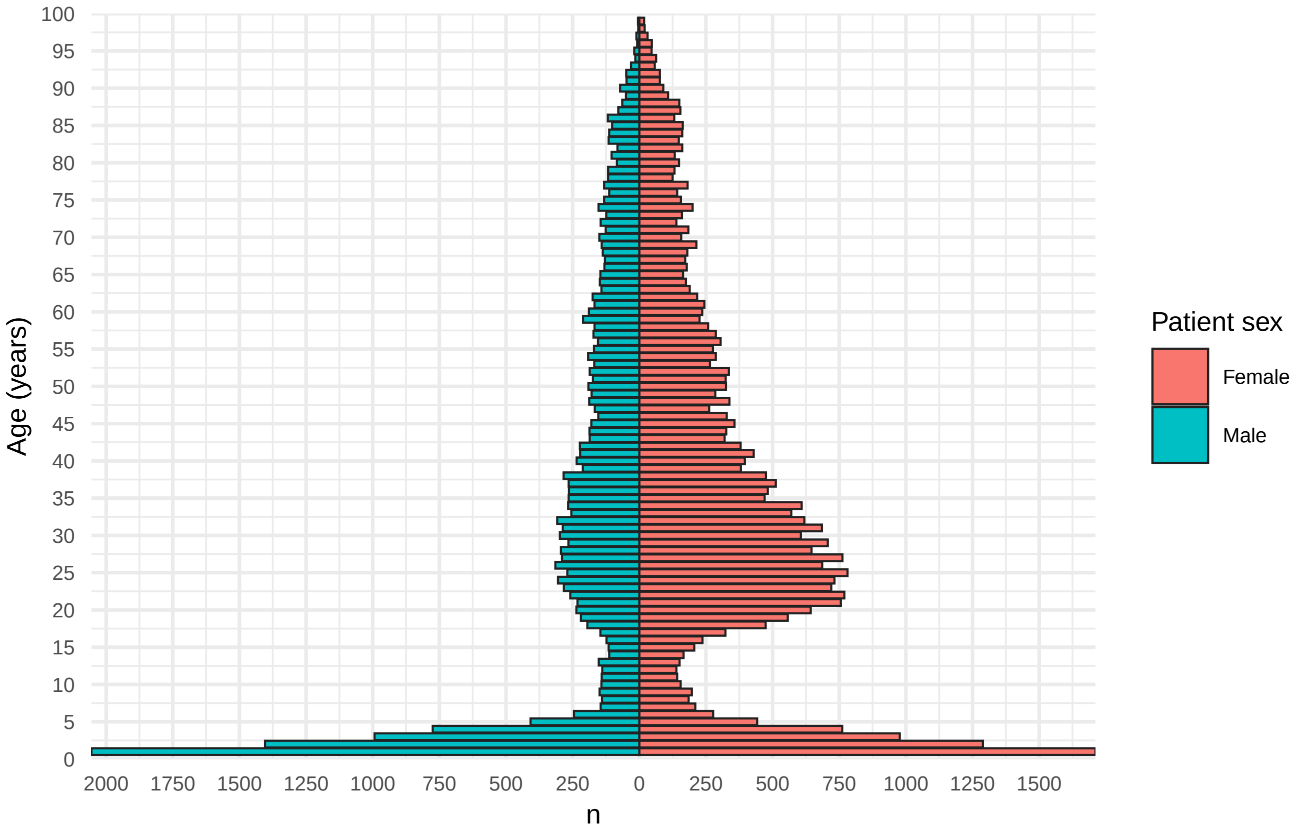


Figure 1: Population pyramid for index 111 calls
