## Supplementary 3 for "An analysis of NHS 111 demand for primary care services"

### Direct booking by 111 call handler

Only GP appointments appeared to be bookable by the 111 call handler based on the data in this cohort, although this was infrequently undertaken and mostly 'in-hours'

| Characteristic | **No**, N = 47,979 | **Yes**, N = 6,037 | **Overall**, N = 54,016 |
|---|---|---|---|
| **Time of index 111 call (N, %)** | | | |
| In-hours | 9,234 (19%) | 4,726 (78%) | 13,960 |
| Out-of-hours | 38,745 (81%) | 1,311 (22%) | 40,056 |
| **Triaged primary care contact timeframe (N, %)** | | | |
| 1hr | 10,014 (21%) | 219 (3.6%) | 10,233 |
| 2hrs | 18,340 (38%) | 1,207 (20%) | 19,547 |
| 6hrs | 11,262 (23%) | 1,262 (21%) | 12,524 |
| >6hrs | 8,363 (17%) | 3,349 (55%) | 11,712 |
| **First service contacted following index 111 call (N, %)** | | | |
| 999 | 667 (1.4%) | 46 (0.8%) | 713 |
| ED | 3,236 (6.7%) | 284 (4.7%) | 3,520 |
| GP | 24,375 (51%) | 1,828 (30%) | 26,203 |
| IP | 420 (0.9%) | 80 (1.3%) | 500 |
| IUC | 2,164 (4.5%) | 231 (3.8%) | 2,395 |
| No further healthcare service contact | 17,117 (36%) | 3,568 (59%) | 20,685 |
