## Supplementary 4 for "An analysis of NHS 111 demand for primary care services"

### Clinical advisor involvement in 111 calls

Clinicians (clinical advisors) are involved in calls in cases where the call handler cannot prioritise the main symptom, the caller response to three NHS Pathways questions is 'unsure', the caller has declared a pre-existing medical condition, refuses the allocated disposition or is deemed complex, and enquiries about medication or medical/surgical procedures. This happened infrequently in this cohort of callers.

| Characteristic | **No**, N = 46,279 | **Yes**, N = 9,823 | **Overall**, N = 56,102 |
|---|---|---|---|
| **Time of index 111 call (N, %)** | | | |
| Out-of-hours | 33,667 (73%) | 7,663 (78%) | 41,330 |
| In-hours | 12,612 (27%) | 2,160 (22%) | 14,772 |
| **Triaged primary care contact timeframe (N, %)** | | | |
| 1hr | 8,241 (18%) | 2,107 (21%) | 10,348 |
| 2hrs | 16,850 (36%) | 2,930 (30%) | 19,780 |
| 6hrs | 10,323 (22%) | 2,571 (26%) | 12,894 |
| >6hrs | 10,865 (23%) | 2,215 (23%) | 13,080 |
| **Triage symptom group (N, %)** | | | |
| Other | 26,600 (57%) | 6,811 (69%) | 33,411 |
| Pain and/or Frequency Passing Urine | 2,970 (6.4%) | 284 (2.9%) | 3,254 |
| Unwell, Under 1 Year Old | 2,273 (4.9%) | 222 (2.3%) | 2,495 |
| Skin, Rash | 1,999 (4.3%) | 441 (4.5%) | 2,440 |
| Earache | 2,216 (4.8%) | 186 (1.9%) | 2,402 |
| Sore Throat or Hoarse Voice | 1,952 (4.2%) | 366 (3.7%) | 2,318 |
| Chest and Upper Back Pain | 1,597 (3.5%) | 511 (5.2%) | 2,108 |
| Vomiting | 1,838 (4.0%) | 218 (2.2%) | 2,056 |
| Lower Back Pain | 1,716 (3.7%) | 232 (2.4%) | 1,948 |
| Cough | 1,579 (3.4%) | 286 (2.9%) | 1,865 |
| Abdominal Pain | 1,539 (3.3%) | 250 (2.5%) | 1,789 |
| **First service contacted following index 111 call (N, %)** | | | |
| GP | 22,178 (48%) | 4,512 (46%) | 26,690 |
| No further healthcare contact | 17,749 (38%) | 4,000 (41%) | 21,749 |
| ED | 3,258 (7.0%) | 545 (5.5%) | 3,803 |
| IUC | 2,100 (4.5%) | 502 (5.1%) | 2,602 |
| 999 | 567 (1.2%) | 172 (1.8%) | 739 |
| IP | 427 (0.9%) | 92 (0.9%) | 519 |
